# Online unsupervised performance-based cognitive testing: a feasible and reliable approach to scalable cognitive phenotyping of Parkinson’s patients

**DOI:** 10.1101/2024.07.03.24309913

**Authors:** Nasri Balit, Sophie Sun, Yilin Zhang, Madeleine Sharp

**Affiliations:** Department of Neurology and Neurosurgery, Montréal Neurological Institute, McGill University, Montréal, Canada

## Abstract

**Introduction:** A better understanding of the heterogeneity in the cognitive and mood symptoms of Parkinson’s disease will require research conducted in large samples of patients. Fully online and remote research assessments present interesting opportunities for scaling up research but the feasibility and reliability of remote and fully unsupervised performance-based cognitive testing in individuals with Parkinson’s disease is unknown. This study aims to establish the feasibility and reliability of this testing modality in Parkinson’s patients.

**Methods:** Sixty-seven Parkinson’s patients and 36 older adults completed two sessions of an at-home, online battery of five cognitive tasks and three self-report questionnaires. Feasibility was established by examining completion rates and data quality. Test-retest reliability was evaluated using the Intraclass Correlation Coefficient (ICC (2,1)).

**Results:** Overall completion rates and data quality were high with few participant exclusions across tasks. With regards to test-retest reliability, intraclass correlation coefficients were quite variable across measures extracted from a task as well as across tasks, but at least one standard measure from each task achieved moderate to good reliability levels. Self-report questionnaires achieved a higher test-retest reliability than cognitive tasks. Feasibility and reliability were similar between Parkinson’s patients and older adults.

**Conclusion:** These results demonstrate that remote and unsupervised testing is a feasible and reliable method of measuring cognition and mood in Parkinson’s patients that achieves levels of test-retest reliability that are comparable to those reported for standard in-person testing.

## Introduction

Although Parkinson’s disease is clinically diagnosed on the basis of motor symptoms, cognitive and mood symptoms are also prevalent and impactful symptoms of the disease [1]. However, the presence, timing and severity of these neuropsychiatric symptoms vary significantly from patient to patient, and little is known about the causes underlying this heterogeneity [2]. Better understanding this heterogeneity will require the evaluation of large samples of Parkinson’s patients with comprehensive assessments of cognitive and mood function. To this end, there has been an increasing interest in turning to fully online and remote research assessments, as this mode of data collection is more easily scalable than in-person research protocols [3]. However, most studies that rely on online assessments, particularly those in clinical populations, such as Parkinson’s patients, include only self-reported measures of symptoms and function rather than performance-based measures, which are necessary to better characterize and quantify the cognitive deficits of Parkinson’s disease [4]. One reason for the omission of cognitive performance tests in large-scale online studies is that the feasibility and reliability of fully remote and unsupervised online cognitive assessments has not yet been established in Parkinson’s disease. This is especially relevant as factors such as computer literacy in older age and disease-related impairments (e.g., motor slowing or mild cognitive impairment) may interfere with the computer-based and unsupervised nature of this mode of data collection [5].

Prior studies of remote, unsupervised cognitive testing have primarily established its reliability in non-clinical adult populations [6–9]. For instance, one study conducted remote unsupervised testing in older adults who completed a battery of tasks assessing visual memory, attention, and executive function on a monthly basis [6]. They found that reliability, measured with the Intraclass Correlation Coefficient (ICC), ranged from 0.50 to 0.76 across tasks, indicating moderate to good reliability, which is in keeping with reliability calculations found in similar studies [7–9].

In clinical populations of patients with neurodegenerative diseases, comparatively little work has been done to establish the test-retest reliability of remote, unsupervised cognitive testing. This likely reflects concerns that, on account of motor and cognitive symptoms which could interfere with data quality, using this mode of testing is not feasible. In Parkinson’s disease, however, there are promising preliminary results. First, with respect to feasibility, a recent study conducted fully online and unsupervised research assessments that consisted of an extensive battery of questionnaires in a large sample of over 20,000 Parkinson’s patients [3]. This study showed good rates of questionnaire completion, with only a 10.1% drop-off rate from the first to the last assessment in the battery [3]. In a follow-up study, Parkinson’s patients within this cohort were shown to be demographically representative to Parkinson’s patients tested in traditional, in-person cohorts [4]. Though these studies did not include any performance-based tasks, which are likely more susceptible to the effects of poor task engagement than self-report questionnaires, the results nonetheless suggest that fully remote and unsupervised interactions with participants are feasible for Parkinson’s disease research. Second, with respect to reliability, one study compared the test-retest reliability of standard in-person testing to supervised virtual testing in Parkinson’s patients, where patients were at home and supervised by a rater on a video call [10]. The results of this study showed that though test-retest reliability between the in-person and virtual administration varied across tasks, many of the tasks achieved at least moderate reliability, which is a level of reliability comparable to that of standard in-person paper-based cognitive testing in Parkinson’s patients [11]. These results are promising but the test-retest reliability of fully remote, unsupervised testing in individuals with Parkinson’s disease remains to be determined prior to large-scale adoption of this mode of data collection.

To address this gap, the objectives of this study were first, to evaluate the feasibility of remote unsupervised cognitive and mood testing by assessing completion rates and data quality, and second, to evaluate the test-retest reliability of the unsupervised tasks and questionnaires. Sixty-seven individuals with Parkinson’s disease and 36 older adults completed two sessions of the online protocol, which consisted of five cognitive tasks targeting working memory, executive function, sustained attention and perceptual decision-making, and three self-report questionnaires assessing mood and cognitive function. Overall, we found that completion rates and data quality were high, with very few participants failing inclusion criteria and attention checks. Although reliability was quite variable across measures, we found that task performance and questionnaire scores achieved at least moderate to good levels of reliability and that reliability was generally similar for Parkinson’s patients and older adult controls. These results suggest that remote and unsupervised testing is a feasible and reliable method of measuring cognition and mood in Parkinson’s patients that achieves levels of test-retest reliability that are comparable to those reported for standard in-person testing.

## Methods

### Participants

We recruited a sample of 70 Parkinson’s patients (PD) and 36 older adults (OA) between the ages of 50-90 through a collaboration with two Parkinson’s patient registries, the Quebec Parkinson’s Network (QPN) and the Canadian Open Parkinson’s Network (C-OPN), which rely on a neurologist-confirmed Parkinson’s disease diagnosis for inclusion in the registry [12]. No additional diagnostic confirmation was conducted in the patients for the purposes of the present study. Older adults were additionally recruited from the community. To capture a sample as representative as possible, we had no exclusion criteria other than age. The 106 participants of the present study were recruited from a larger sample of 223 participants (144 PD and 74 OA) who had completed a more extensive initial online assessment (the results of which will be reported separately), and who agreed to complete testing at a second timepoint after an interval that ranged from 21 to 135 days. The participants included in the present study are those who completed, at minimum, one task of the second assessment (Time 2). Three Parkinson’s patients were excluded because the interval between their assessments was either less than 21 days or more than 135 days. The final sample included 67 Parkinson’s patients and 36 older adults (Table 1). Of the 67 Parkinson’s patients and 36 older adults, 8 Parkinson’s patients and 2 older adults (9.71% of total participants) had only partial data for Time 2. All participants were entered in a draw to win one of ten $100 gift cards. All procedures were evaluated and approved by the McGill University Health Centre (MUHC) Research Ethics Board.

**Table 1:**
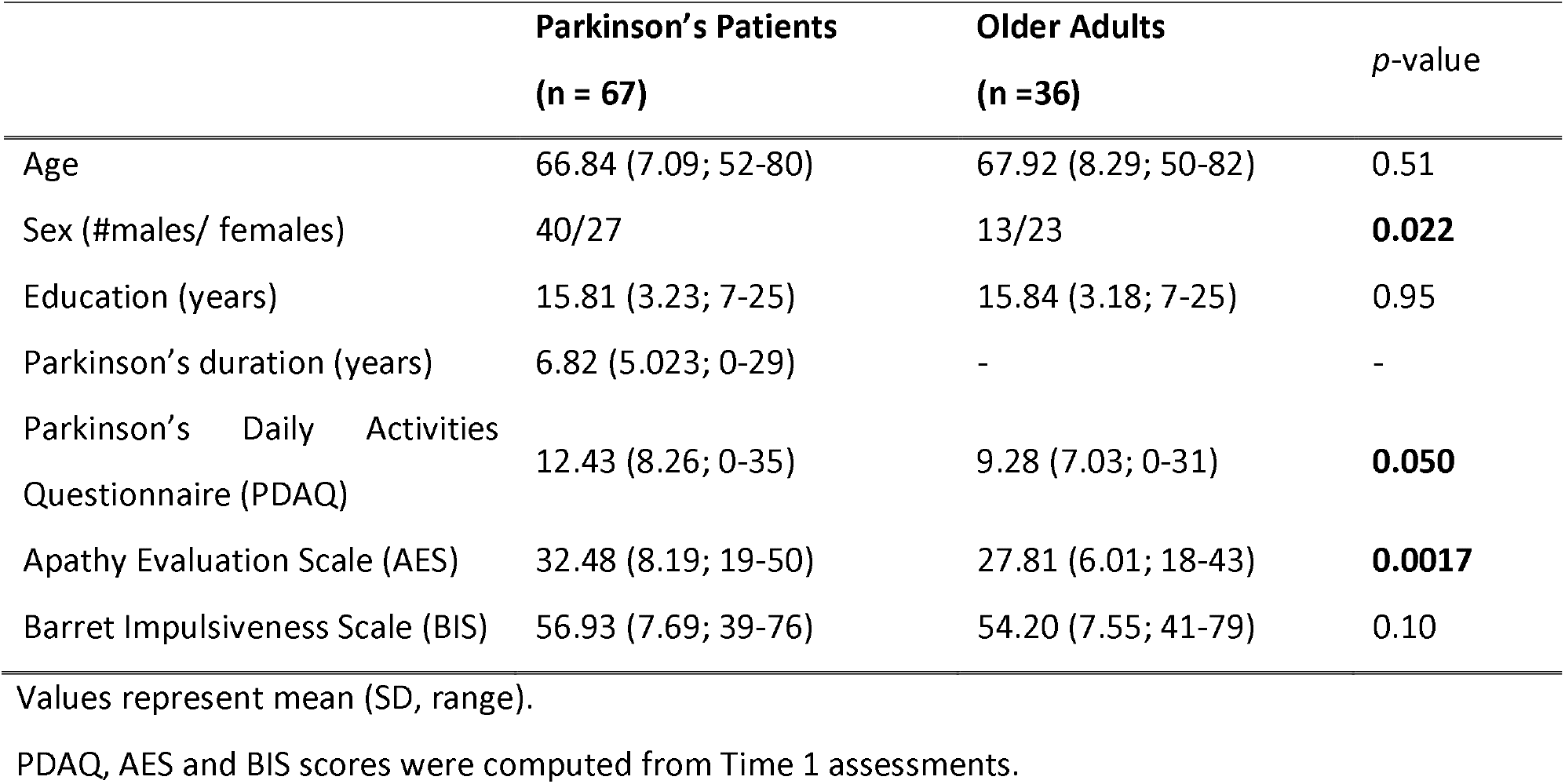
Participant characteristics.

|  | **Parkinson's Patients**<br>**(n = 67)** | **Older Adults**<br>**(n =36)** | *p*-value |
| --- | --- | --- | --- |
| Age | 66.84 (7.09; 52-80) | 67.92 (8.29; 50-82) | 0.51 |
| Sex (#males/ females) | 40/27 | 13/23 | **0.022** |
| Education (years) | 15.81 (3.23; 7-25) | 15.84 (3.18; 7-25) | 0.95 |
| Parkinson's duration (years) | 6.82 (5.023; 0-29) | - | - |
| Parkinson's Daily Activities<br>Questionnaire (PDAQ) | 12.43 (8.26; 0-35) | 9.28 (7.03; 0-31) | **0.050** |
| Apathy Evaluation Scale (AES) | 32.48 (8.19; 19-50) | 27.81 (6.01; 18-43) | **0.0017** |
| Barret Impulsiveness Scale (BIS) | 56.93 (7.69; 39-76) | 54.20 (7.55; 41-79) | 0.10 |

Values represent mean (SD, range).
PDAQ, AES and BIS scores were computed from Time 1 assessments.

### Procedure

Potential participants, i.e. those who had completed the Time 1 online assessment, were emailed with information on our study and a link to the consent form. The Time 1 assessment was more extensive and contained a battery of ten cognitive tasks and six mood questionnaires. Of these, to enhance recruitment and keep the burden on participants to a minimum, only five cognitive tasks and three mood questionnaires were selected for inclusion in the reliability assessment (Time 2), and only this subset will be described here. Expected total time required to complete these tests was 45 minutes and participants could pause between (but not during) tasks. All testing was conducted fully remotely, from personal desktop or laptop computers (smartphones and tablets were not allowed), and there was no interaction with the research team during testing.

### Cognitive tasks & mood questionnaires

The cognitive tasks included in the protocol were selected because they are tests of domains of cognition affected early in Parkinson’s disease [13]. We included computerized versions of Trail Making [14], Digit Span [15], Color-word Stroop [16], and Sustained Attention to Response (SART) [17] tasks. We also included a signal-detection task [18], which is not a standard measure of cognition in Parkinson’s patients but was included because it is considerably longer (15 minutes) and therefore could be more susceptible to the effects of remote unsupervised testing on task engagement. The self-report questionnaires assessed apathy (Apathy Evaluation Scale; AES) [19], impulsivity (Barrett Impulsiveness Scale-11; BIS-11) [20], and everyday cognitive function (Parkinson’s Daily Activities Questionnaire; PDAQ) [21].

Each task started with a set of detailed on-screen instructions, followed by a short practice, and then a review of the key instructions before the initiation of the main phase of the task. In addition, each questionnaire embedded an extra item that was a prompt to check for attention (e.g., “Select ‘Not likely’. This is simply to ensure that you are paying attention”), a common practice in online behavioural research [22]. More detailed information on our administration of the tasks and questionnaires can be found in the Supplementary Methods.

### Assessing data quality to establish feasibility

We used data quality as our main measure of feasibility, which we defined as the proportion of participants who were excluded because they failed minimum data quality checks.

#### Cognitive tasks

We excluded participants who performed two standard deviations below the mean on one timepoint (of a given task) and 2 standard deviations above the mean on the other timepoint of that same task. We assumed that such a large variation in performance suggest that external circumstances leading to poor task engagement may have affected their performance on one of the two testing instances. Additionally, in the case of the Stroop, signal detection and SART tasks, we excluded participants who either did not input a response to any of their trials or responded using the same key for each trial at either of the 2 timepoints. Proportions of participants excluded were computed for each task.

#### Questionnaires

We computed the proportion of participants excluded for failing the attention check.

### Data Analysis

Welch two sample *t* tests were used to evaluate group differences and intraclass correlation coefficients (ICC) were calculated to assess test-retest reliability, separately for the Parkinson’s patients and older adults. According to the guideline outlined in Koo & Li [23], a two-way mixed effects model for consistency and with a single rater was used (ICC (2,1)). The correlation coefficient was interpreted according to the following ranges, with ICCs <0.5, 0.5-0.75, 0.75-0.90 and >0.90 indicating poor, moderate, good and excellent reliability respectively [23]. Paired t tests were used to determine differences in performance between the two timepoints and evaluate the presence of practice effects. The critical p value in all analyses was set to 0.05. All analyses were conducted in R version 4.2.3 [24].

## Results

### Assessment of data quality

For self-report questionnaires, an average of 3.33 (4.97%) Parkinson’s patients (PD) and 0.33 (0.92%) Older Adults (OA) were excluded for each questionnaire after failing the ‘attention check’ at either timepoint. For the cognitive tasks, only Trail Making, Stroop and SART had participant exclusions based on our previously defined criteria. In Trail Making, six PD (10.17%) and 4 OA (11.43%) were excluded from part A and 2 PD (3.39%) and 2 OA (5.71%) were excluded from part B. In Stroop, 4 PD (6.06%) and 1 OA (2.86%) were excluded. In SART, 2 PD (3.23%) and 1 OA (2.86%) were excluded from both conditions. Ten participants (9.71%), of which 8 were PD (11.94%) and 2 were OA (5.56%), dropped out before completing Time 2.

### Comparison of performance between Parkinson’s patients and older adults

As shown in **Table 1**, both patients and older adults reported similar levels of mood symptoms except that PD patients reported a higher level of apathy symptoms (AES: t(90.69)=3.24, *p*=0.0017) as well as a higher impairment of daily activities (PDAQ: t(83.11)=1.99, *p*=0.0497).

With respect to the cognitive tasks, there were no statistically significant differences in performance between the PD and OA on the Digit Span, Stroop and SART tasks (all *p*s>0.05; **Table 2**). Parkinson’s patients were slower on the Trail making Part A (t(81.83)=3.74, *p*<0.001) and on the Trail making Part B (t(71.47)=3.18, *p*=0.0022). On the Signal Detection task, Parkinson’s patients were slower (t(94.95)=2.25, *p*=0.027); and had lower discriminability (t(81.52)=4.75, *p*<0.001). Additional secondary outcome measures that can be derived from each task, such as accuracy, also showed similar performance between groups (**Supplementary Table S1**).

**Table 2:**
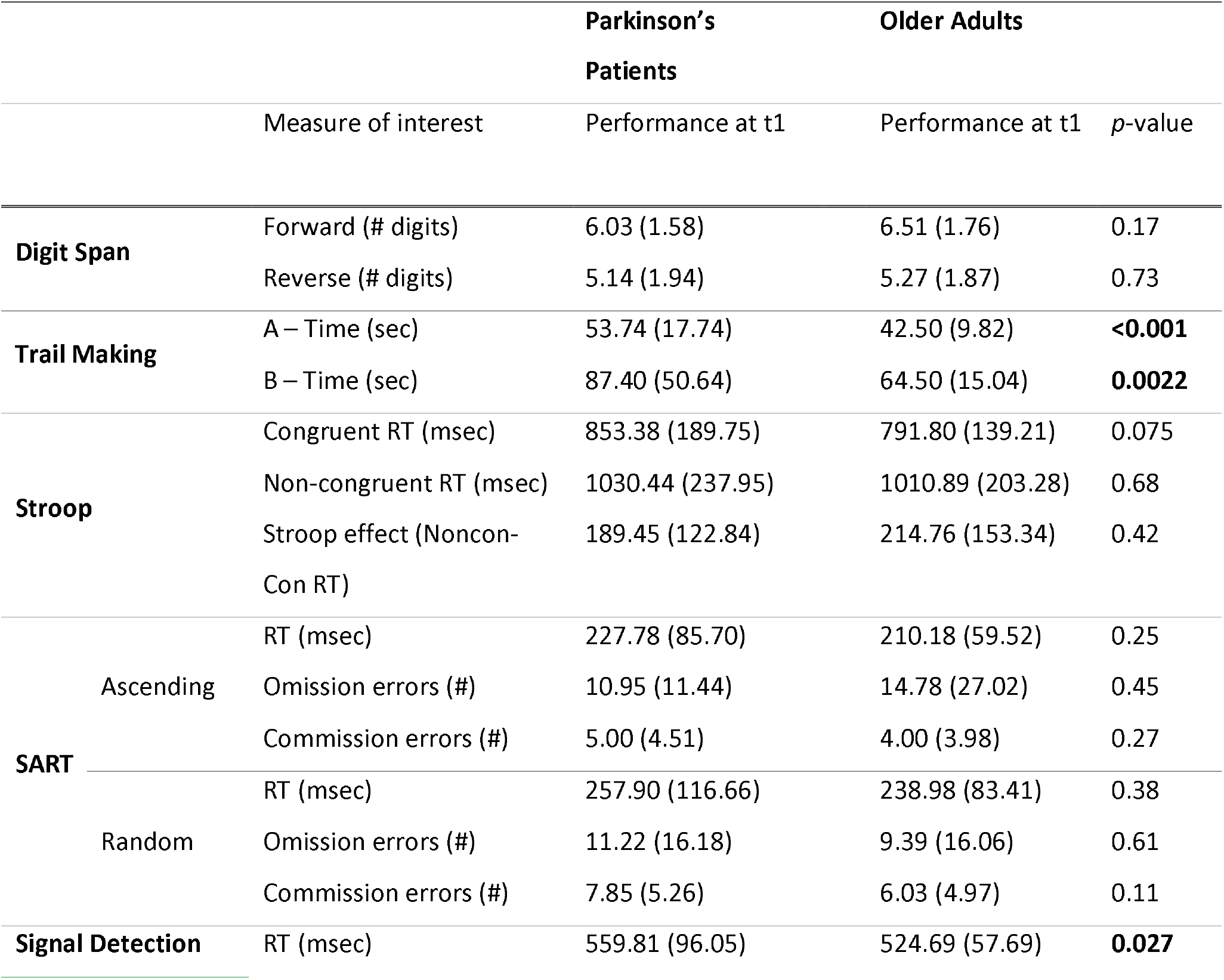
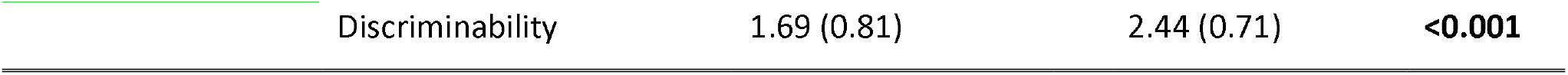
Performance on cognitive tasks.

### Test-retest reliability of cognitive tasks

For each of the tasks, at least one of the outcome measures extracted demonstrated moderate to good reliability in both Parkinson’s patients and older adults, but there was notable variability between tasks and even between different measures extracted from different conditions of the same task (**Table 3**). For instance, in Parkinson’s patients, forward Digit Span demonstrated an ICC of 0.49 and backward Digit Span had an ICC of 0.71, whereas the ICCs for parts A and B of Trail Making were very similar. In the case of the Stoop task, the derived measure (i.e. ‘Stroop effect’) had very poor reliability (ICC=0.035) whereas the reliability for average response times on both congruent and non-congruent trials were higher (part A: ICC=0.73; part B: ICC=0.41). In the case of the SART, the time-dependent outcome measures (Ascending-RT: ICC=0.73; Descending-RT: ICC=0.74) had higher reliability than the accuracy-based outcome measures, for which ICCs ranged from 0.36 to 0.60. For the Signal Detection task, both the raw response time-based outcome measure (RT: ICC=0.80) and the derived outcome measure (Discriminability: ICC=0.78) demonstrated good reliability. Reliability of alternative outcome measures were in a similar range (**Supplementary Table S4**). Similar ranges of ICCs across tasks were also found in older adults (**Table 3** and **Supplementary Table S4**).

**Table 3:**
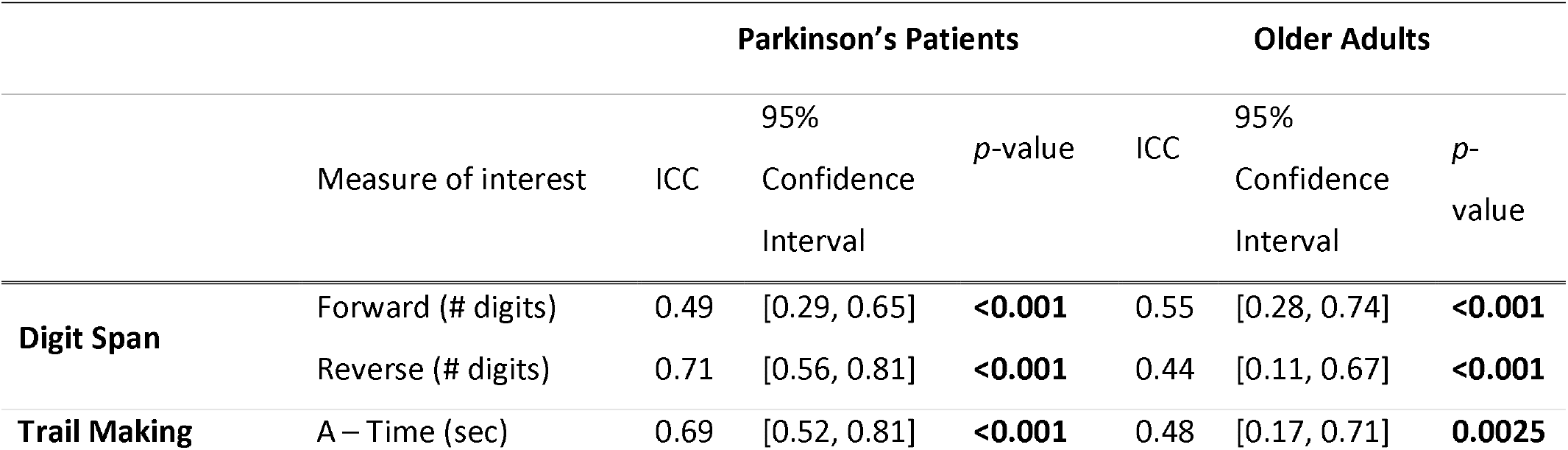
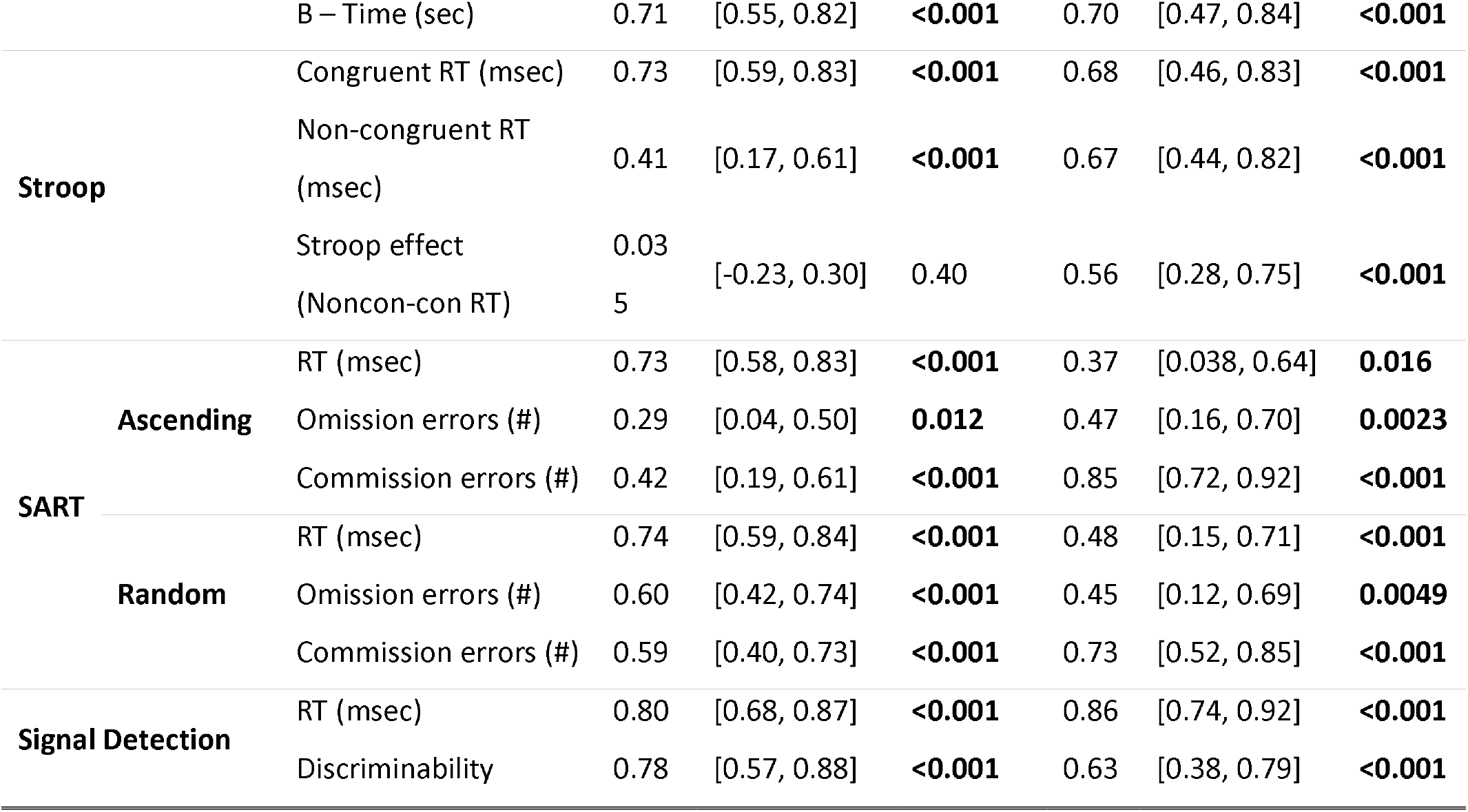
Test-retest reliability of cognitive tasks.

### Test-retest reliability of self-report questionnaires

As shown in **Table 4**, test-retest reliability for the mood and cognitive function questionnaires were good in both the PD patients (ICC range: 0.77-0.87) and the OAs, with the exception of the PDAQ which was in the moderate range in OAs (ICC=0.56).

**Table 4:** Test-retest reliability of mood questionnaires.

| Questionnaire | Parkinson's Patients |  |  | Older Adults |  |  |
| --- | --- | --- | --- | --- | --- | --- |
|  | ICC | p-value | 95% Confidence Interval | ICC | p-value | 95% Confidence Interval |
| Parkinson's Daily Activities Questionnaire (PDAQ) | 0.87 | **<0.001** | [0.78, 0.92] | 0.56 | **<0.001** | [0.25, 0.76] |
| Apathy Evaluation Scale (AES) | 0.77 | **<0.001** | [0.65, 0.86] | 0.86 | **<0.001** | [0.74, 0.92] |
| Barratt Impulsiveness Scale (BIS) | 0.78 | **<0.001** | [0.65, 0.86] | 0.81 | **<0.001** | [0.62, 0.90] |

### Practice effects

In Parkinson’s patients, no significant improvement in performance was observed across the two timepoints (*p*s>0.05; **Supplementary Table S2**). The only exception was a speeding of responses on the ascending and random condition of the SART (Asc mean change=-15.96ms, *p*=0.048; Ran mean change = −23.44, *p*=0.018) as well as an increased discriminability in the signal detection task (mean change=0.28, *p*=<0.001) in PDs. In OAs, a speeding of responses in the random condition of the SART was observed (mean change=-39.42ms, *p*=0.0087), as well as an increased span in the reverse Digit Span condition (mean change=1.00, *p*=<0.001). Practice effects are also reported for alternative measures of interest in **Supplementary Table S3**.

## Discussion

The goals of this study were to determine the feasibility and test-retest reliability of online unsupervised performance-based cognitive tasks and mood questionnaires in Parkinson’s patients. We found that data quality was high across all tasks and questionnaires, resulting in very few exclusions, and that test-retest reliability was at least moderate to good for at least one measure of interest from each task and was similar in both Parkinson’s patients and older adults. These results suggest that online and fully unsupervised measurements of mood and cognition are feasible and reliable in Parkinson’s patients and that this mode of testing can be incorporated into online clinical research studies.

We found moderate or higher test-retest reliability (ICCs >0.5) for at least one of the standard outcome measures extracted from each performance-based cognitive task in both Parkinson’s patients and older adults. These results build on recent work, conducted primarily in older adults, that has shown that test-retest reliability of remote, unsupervised administrations of cognitive tasks, including tasks of attention, working memory and executive function is in the moderate to good range [8,9,25]. For instance, one study in older adults examining test-retest reliability of a remote administration of the Trail Making and Stroop tasks, two tasks also included in our battery, found ICC values of 0.5-0.74 [6]. More importantly, however, the level of reliability we found across tasks was comparable to that reported for more traditional supervised, in-person cognitive testing, which remains the ‘gold standard’ of cognitive testing. For instance, in a study assessing the test-retest reliability of an in-person battery of ten cognitive tasks administered to a sample of Parkinson’s patients, reliability values ranged from 0.40-0.75, calculated via weighted Cohen’s kappa [11]. Our results therefore suggest that despite the early mild cognitive deficits that can be present in patients with Parkinson’s disease, and despite the presence of motor deficits, both of which could interfere with remote cognitive testing, the effect of having Parkinson’s disease does not disproportionately impact the reliability of remote cognitive testing in Parkinson’s patients when compared to non-clinical older adult populations. This suggests that unsupervised remote cognitive testing of Parkinson’s patients can be considered as a reliable alternative to paper-based, supervised assessments in the design of clinical research protocols.

Test-retest reliability of the self-report questionnaires assessing apathy, implusivity and daily living ability ranged from 0.77 to 0.87 indicating good reliability. This is consistent with the range of reliability levels reported in the original, paper-based administrations of these questionnaires. Reported test-retest reliability for the Apathy Evaluation Scale and Barratt Impulsiveness scale was 0.76-0.94 [19] and 0.83 [26], respectively. For the Parkinson’s Disease Activities Questionnaire, the test-retest reliability of the shorter, 15 item version used in our study has not been assessed, however the original 50 item version of the questionnaire was demonstrated to have a high ICC of 0.97 [27].

As expected, the test-retest reliability of the self-report questionnaires was, on average, higher than that of the cognitive measures. This aligns with one other study providing a comparison of the reliability of performance-based cognitive tasks and self-report questionnaires [28]. The authors suggested that the lower test-retest reliability of performance-based tasks might reflect the fact that cognitive tasks, in contrast to mood questionnaires, are typically designed to maximize between group differences, which results in comparatively lower between-subject variability. Because test-retest reliability is computed as the ratio of between-subject variance over total variance, the resulting ICC of cognitive tasks is lower [28,29]. Given the increasing interest in the field of Parkinson’s disease to relate individual differences in cognitive function to underlying features of the neurodegenerative process, clinical research protocols aimed at cognitive phenotyping will have to ensure the selection of performance-based measaures with good psychometic properties.

We found that data quality, across both the performance-based cognitive tasks and the self-report questionnaires was high in both the Parkinson’s patients and the older adults. Fewer than 5% of participants, on average, were excluded for failing attention checks on the questionnaires. Similarly, less than 4% of PD and OA participants were excluded based on data quality, on average, for each cognitive task. Additionally, over 90% of participants completed the full protocol. Although the feasibility of remote, unsupervised performance-based cognitive testing in Parkinson’s patients has not explicitly been previously explored, other studies have demonstrated the feasibility of using self-report questionnaires for large-scale, remote, research in Parkinson’s patients. For instance, the Fox Insight study, a large online-only study, has successfully recruited a very large cohort of individuals with and without Parkinson’s disease to complete longitudinal assessments and has shown that self-report assessments obtained online are comparable to those obtained during in-person assessments [4,5]. Our results suggest that the added complexity of cognitive testing does no significantly hinder the feasibilty of the remote, unsupervised testing modality but future work is need in patients with more advanced disease and a higher burden of cognitive deficits to ensure this mode of testing can be expanded to a more representative patient population, and in particular, to ensure that large-scale online testing can be leveraged to better understand the progression to mild cognitive impairment and to dementia.

Another factor potentially limiting the utility of performance-based cognitive tasks is the presence of practice effects. It is conceivable that such effects might be accentuated in online computer-based research given that participants’ performance might benefit from prior exposure to the computer-based and at-home setting (e.g., learning to minimize distactions, gaining familiarity with typical key responses, etc.). Overall, however, we did not find a pattern of performance change over time consistent with practice. Performance was sligtly worse on two tasks, and slightly better (faster responses) for one of the measures of a different task. Given the inconsistent changes, it is unlikely that this represents meaningful disease-related changes. Furthermore, given that the interval between testing sessions (20 to 135 days) was much shorter than the typical interval between assessments in longitudinal research, even possible small practice effects would likely abate over longer intervals.

One limitation of our study is that our sample is likely not representative of the greater Parkinson’s patient population. Though we successfully recruited patients with a range of disease duration (mean duration 6.82 years), as well as a higher proportion of females (40% female) than is typically reported [30], the education level was high and perhaps most telling, the cognitive performance and mood differences between the patients and older adults were minimal. This suggests a selection bias towards patients with earlier and/or milder disease. Though the inclusion of patients with significant cognitive impairment might be beyond the scope of online research, we think that certain modifications, such as shortening the protocol, ‘gamifying’ the cognitive tasks, and developing smartphone-compatible assessments could help bolster recruitment from a more diverse population of patients but future work engaging with patients is needed in order to identify and address the full range of barriers to participation.

In summary, these results indicate that online and unsupervised performance-based cognitive testing and self-report-based mood testing conducted in Parkinson’s patients is feasible and produces levels of reliability that are comparable to that of standard in-person testing. There is already evidence that leveraging remote online testing results in enrolment of much larger samples of Parkinson’s patients than would otherwise be possible with in-person testing. Our results suggest that more in depth cognitive and neuropsychiatric phenotyping is also possible on this scale, which is an important step towards designing research studies that are aimed at identifying the potential mechanisms underlying the heterogeneity in Parkinson’s disease.

## Data Availability

All data produced in the present study are available upon reasonable request to the authors.

## Acknowledgements

We would like to thank the participants for their time.

## Author contributions

**Nasri Balit:** Formal analysis, Investigation, Methodology, Software, Visualization, Writing – original draft. **Sophie Sun:** Investigation, Methodology, Writing – review & editing. **Yilin Zhang:** Investigation, Methodology, Writing – review & editing. **Madeleine Sharp:** Conceptualization, Methodology, Project administration, Resources, Supervision, Writing – review & editing.

## Data and materials availability

All cognitive task code is openly available at https://github.com/cognitive-neuroscience/neuron and all analysis scripts are available at https://osf.io/vdw5h/. De-identified raw data is available upon request.

## References

[1] C.H. Williams-Gray, J.R. Evans, A. Goris, T. Foltynie, M. Ban, T.W. Robbins, C. Brayne, B.S. Kolachana, D.R. Weinberger, S.J. Sawcer, R.A. Barker, The distinct cognitive syndromes of Parkinson’s disease: 5 year follow-up of the CamPaIGN cohort, Brain J. Neurol. 132 (2009) 2958–2969. 10.1093/brain/awp245.

[2] T. Foltynie, C. Brayne, R.A. Barker, The heterogeneity of idiopathic Parkinson’s disease, J. Neurol. 249 (2002) 138–145. 10.1007/pl00007856.

[3] L. Smolensky, N. Amondikar, K. Crawford, S. Neu, C.M. Kopil, M. Daeschler, L. Riley, E. Brown, A.W. Toga, C. Tanner, Fox Insight collects online, longitudinal patient-reported outcomes and genetic data on Parkinson’s disease, Sci. Data 7 (2020) 67. 10.1038/s41597-020-0401-2.

[4] L.M. Chahine, I. Chin, C. Caspell-Garcia, D.G. Standaert, E. Brown, L. Smolensky, V. Arnedo, D. Daeschler, L. Riley, M. Korell, R. Dobkin, N. Amondikar, S. Gradinscak, I. Shoulson, M. Dean, K. Kwok, P. Cannon, K. Marek, C. Kopil, C.M. Tanner, C. Marras, Comparison of an Online-Only Parkinson’s Disease Research Cohort to Cohorts Assessed In Person, J. Park. Dis. 10 (n.d.) 677–691. 10.3233/JPD-191808.

[5] C. Malinowsky, A. Kottorp, A. Wallin, A. Nordlund, E. Björklund, I. Melin, A. Pernevik, L. Rosenberg, L. Nygård, Differences in the use of everyday technology among persons with MCI, SCI and older adults without known cognitive impairment, Int. Psychogeriatr. 29 (2017) 1193–1200. 10.1017/S1041610217000643.

[6] K.E. Dorociak, N. Mattek, J. Lee, M.I. Leese, N. Bouranis, D. Imtiaz, B.M. Doane, J.P.K. Bernstein, J.A. Kaye, A.M. Hughes, The Survey for Memory, Attention, and Reaction Time (SMART): Development and Validation of a Brief Web-Based Measure of Cognition for Older Adults, Gerontology 67 (2021) 740–752. 10.1159/000514871.

[7] N.A. Kochan, M. Heffernan, M. Valenzuela, P.S. Sachdev, B.C.P. Lam, M. Fiatarone Singh, K.J. Anstey, T. Chau, H. Brodaty, Reliability, Validity, and User-Experience of Remote Unsupervised Computerized Neuropsychological Assessments in Community-Living 55-to 75-Year-Olds, J. Alzheimers Dis. JAD 90 (2022) 1629–1645. 10.3233/JAD-220665.

[8] J.R. Myers, J.M. Glenn, E.N. Madero, J. Anderson, R. Mak-McCully, M. Gray, J.L. Gills, J.E. Harrison, Asynchronous Remote Assessment for Cognitive Impairment: Reliability Verification of the Neurotrack Cognitive Battery, JMIR Form. Res. 6 (2022) e34237. 10.2196/34237.

[9] N.H. Stricker, J.L. Stricker, A.J. Karstens, J.R. Geske, J.A. Fields, J. Hassenstab, C.G. Schwarz, N. Tosakulwong, H.J. Wiste, C.R. Jack, K. Kantarci, M.M. Mielke, A novel computer adaptive word list memory test optimized for remote assessment: Psychometric properties and associations with neurodegenerative biomarkers in older women without dementia, Alzheimers Dement. Amst. Neth. 14 (2022) e12299. 10.1002/dad2.12299.

[10] J. Gallagher, E. Mamikonyan, S.X. Xie, B. Tran, S. Shaw, D. Weintraub, Validating virtual administration of neuropsychological testing in Parkinson disease: a pilot study, Sci. Rep. 13 (2023) 16243. 10.1038/s41598-023-42934-0.

[11] J. Marinus, M. Visser, N.A. Verwey, F.R.J. Verhey, H. a. M. Middelkoop, A.M. Stiggelbout, J.J. van Hilten, Assessment of cognition in Parkinson’s disease, Neurology 61 (2003) 1222–1228. 10.1212/01.wnl.0000091864.39702.1c.

[12] Z. Gan-Or, T. Rao, E. Leveille, C. Degroot, S. Chouinard, F. Cicchetti, A. Dagher, S. Das, A. Desautels, J. Drouin-Ouellet, T. Durcan, J.-F. Gagnon, A. Genge, J. Karamchandani, A.-L. Lafontaine, S.L.W. Sun, M. Langlois, M. Levesque, C. Melmed, M. Panisset, M. Parent, J.-B. Poline, R.B. Postuma, E. Pourcher, G.A. Rouleau, M. Sharp, O. Monchi, N. Dupré, E.A. Fon, The Quebec Parkinson Network: A Researcher-Patient Matching Platform and Multimodal Biorepository, J. Park. Dis. 10 (2020) 301–313. 10.3233/JPD-191775.

[13] D. Weintraub, T. Simuni, C. Caspell-Garcia, C. Coffey, S. Lasch, A. Siderowf, D. Aarsland, P. Barone, D. Burn, L.M. Chahine, J. Eberling, A.J. Espay, E.D. Foster, J.B. Leverenz, I. Litvan, I. Richard, M.D. Troyer, K.A. Hawkins, the P.P.M. Initiative, Cognitive performance and neuropsychiatric symptoms in early, untreated Parkinson’s disease, Mov. Disord. 30 (2015) 919–927. 10.1002/mds.26170.

[14] Validity of the Trail Making Test as an Indicator of Organic Brain Damage - Ralph M. Reitan, 1958, (n.d.). https://journals.sagepub.com/doi/abs/10.2466/pms.1958.8.3.271 (accessed June 13, 2024).

[15] D. Wechsler, H. Kodama, Wechsler intelligence scale for children, Psychological corporation New York, 1949.

[16] J.R. Stroop, Studies of interference in serial verbal reactions, J. Exp. Psychol. 18 (1935) 643–662. 10.1037/h0054651.

[17] I.H. Robertson, T. Manly, J. Andrade, B.T. Baddeley, J. Yiend, ‘Oops!’: Performance correlates of everyday attentional failures in traumatic brain injured and normal subjects, Neuropsychologia 35 (1997) 747–758. 10.1016/S0028-3932(97)00015-8.

[18] D.A. Pizzagalli, A.L. Jahn, J.P. O’Shea, Toward an objective characterization of an anhedonic phenotype: a signal-detection approach, Biol. Psychiatry 57 (2005) 319–327. 10.1016/j.biopsych.2004.11.026.

[19] R.S. Marin, R.C. Biedrzycki, S. Firinciogullari, Reliability and validity of the Apathy Evaluation Scale, Psychiatry Res. 38 (1991) 143–162. 10.1016/0165-1781(91)90040-v.

[20] M. Luengo, M. Carrillo-De-La-Pena, J. Otero, The components of impulsiveness: A comparison of the I. 7 Impulsiveness Questionnaire and the Barratt Impulsiveness Scale, Personal. Individ. Differ. 12 (1991) 657–667.

[21] L. Brennan, A. Siderowf, J.D. Rubright, J. Rick, N. Dahodwala, J.E. Duda, H. Hurtig, M. Stern, S.X. Xie, L. Rennert, J. Karlawish, J.A. Shea, J.Q. Trojanowski, D. Weintraub, The Penn Parkinson’s Daily Activities Questionnaire-15: Psychometric properties of a brief assessment of cognitive instrumental activities of daily living in Parkinson’s disease, Parkinsonism Relat. Disord. 25 (2016) 21–26. 10.1016/j.parkreldis.2016.02.020.

[22] B.D. Douglas, P.J. Ewell, M. Brauer, Data quality in online human-subjects research: Comparisons between MTurk, Prolific, CloudResearch, Qualtrics, and SONA, PloS One 18 (2023) e0279720. 10.1371/journal.pone.0279720.

[23] T.K. Koo, M.Y. Li, A Guideline of Selecting and Reporting Intraclass Correlation Coefficients for Reliability Research, J. Chiropr. Med. 15 (2016) 155–163. 10.1016/j.jcm.2016.02.012.

[24] R.C. Team, R: A language and environment for statistical computing. R Foundation for Statistical Computing, No Title (2013).

[25] S. Belleville, A.A. LaPlume, R. Purkart, Web-based cognitive assessment in older adults: Where do we stand?, Curr. Opin. Neurol. 36 (2023) 491–497. 10.1097/WCO.0000000000001192.

[26] C.W. Mathias, M.S. Stanford, Y. Liang, M. Goros, N.E. Charles, A.H. Sheftall, J. Mullen, N. Hill-Kapturczak, A. Acheson, R.L. Olvera, D.M. Dougherty, A Test of the Psychometric Characteristics of the BIS-Brief Among Three Groups of Youth, Psychol. Assess. 30 (2018) 847–856. 10.1037/pas0000531.

[27] L. Brennan, A. Siderowf, J.D. Rubright, J. Rick, N. Dahodwala, J.E. Duda, H. Hurtig, M. Stern, S.X. Xie, L. Rennert, J. Karlawish, J.A. Shea, J.Q. Trojanowski, D. Weintraub, Development and Initial Testing of the Penn Parkinson’s Daily Activities Questionnaire, Mov. Disord. Off. J. Mov. Disord. Soc. 31 (2016) 126–134. 10.1002/mds.26339.

[28] A.Z. Enkavi, I.W. Eisenberg, P.G. Bissett, G.L. Mazza, D.P. MacKinnon, L.A. Marsch, R.A. Poldrack, Large-scale analysis of test-retest reliabilities of self-regulation measures, Proc. Natl. Acad. Sci. U. S. A. 116 (2019) 5472–5477. 10.1073/pnas.1818430116.

[29] C. Hedge, G. Powell, P. Sumner, The reliability paradox: Why robust cognitive tasks do not produce reliable individual differences, Behav. Res. Methods 50 (2018) 1166–1186. 10.3758/s13428-017-0935-1.

[30] K. Marek, D. Jennings, S. Lasch, A. Siderowf, C. Tanner, T. Simuni, C. Coffey, K. Kieburtz, E. Flagg, S. Chowdhury, W. Poewe, B. Mollenhauer, P.-E. Klinik, T. Sherer, M. Frasier, C. Meunier, A. Rudolph, C. Casaceli, J. Seibyl, S. Mendick, N. Schuff, Y. Zhang, A. Toga, K. Crawford, A. Ansbach, P.D. Blasio, M. Piovella, J. Trojanowski, L. Shaw, A. Singleton, K. Hawkins, J. Eberling, D. Brooks, D. Russell, L. Leary, S. Factor, B. Sommerfeld, P. Hogarth, E. Pighetti, K. Williams, D. Standaert, S. Guthrie, R. Hauser, H. Delgado, J. Jankovic, C. Hunter, M. Stern, B. Tran, J. Leverenz, M. Baca, S. Frank, C.-A. Thomas, I. Richard, C. Deeley, L. Rees, F. Sprenger, E. Lang, H. Shill, S. Obradov, H. Fernandez, A. Winters, D. Berg, K. Gauss, D. Galasko, D. Fontaine, Z. Mari, M. Gerstenhaber, D. Brooks, S. Malloy, P. Barone, K. Longo, T. Comery, B. Ravina, I. Grachev, K. Gallagher, M. Collins, K.L. Widnell, S. Ostrowizki, P. Fontoura, T. Ho, J. Luthman, M. van der Brug, A.D. Reith, P. Taylor, The Parkinson Progression Marker Initiative (PPMI), Prog. Neurobiol. 95 (2011) 629–635. 10.1016/j.pneurobio.2011.09.005.

