## Supplement for "Online unsupervised performance-based cognitive testing: a feasible and reliable approach to scalable cognitive phenotyping of Parkinson’s patients"

**Supplementary Methods**

**Cognitive tasks**

*Trail Making task, Parts A & B:* As with the standard paper-based version [14], Part A consisted of 25 numbers (from 1 to 25) scattered on the screen. Participants were instructed to click on the numbers in ascending order with their mouse or trackpad as fast as possible. In Part B, 13 numbers (from 1 to 13) and 12 letters (from A to L) were similarly scattered on the screen. Participants were instructed to click in alternating ascending order on a number then on a letter as fast as possible (i.e., 1-A-2-B etc.). Maximum allocated time to complete was four minutes for both parts. The primary measure of interest was the time taken to complete Parts A and B.

*Digit Span task:* Both Forward and Reverse Digit Span tasks were administered [15]. The Forward span task consisted of a series of digits (1-9) appearing one at a time on the screen for 1000ms each, with a fixation cross appearing for 500ms between each digit. Once all the digits appeared, participants were instructed to input the digits in the same order by clicking an on-screen number-pad using their mouse or trackpad. There were seven levels of difficulty, starting with a three-digit sequence and increasing to a maximum of a nine-digit sequence, with two trials per level (using two different digit sequences). The Reverse span task followed the same procedure, but the starting sequence was only two digits and increased to a maximum of an eight-digit sequence. Participants had 30 seconds to input their response on each trial. If participants answered one of the two trials correctly, they proceeded to the next level immediately. The measures of interest were the maximum forward and reverse span.

*Color-word Stroop task:* Similar to the Interference condition of the Stroop task [16], a series of color names (‘red’, ‘blue’, ‘green’) were presented on screen either in the same font color as the word (congruent trials, 90/120 trials) or in one of the other two colors (noncongruent trials, 30/120). Participant were instructed to indicate the color of the font using one of three keyboard keys (‘1’ for red, ‘2’ for blue and ‘3’ for green). Maximum response time allowed on each trial was 2000ms. The measures of interest for this task were the average response time (RT) for correct trials on congruent and noncongruent trials, and the Stroop effect (noncongruent RT minus congruent RT).

*Sustained Attention to Response task (SART):* Participants viewed a sequence of digits presented very briefly on the screen one by one (digits 1 to 9) and were instructed to press the left arrow key as fast as possible following the presentation of each number (Go trials), with the exception that when the digit ‘3’ was presented, they had to withhold their response (NoGo trials). There were two blocks, the order of which was counterbalanced across participants: an Ascending condition, where the digits appeared in ascending order from 1 to 9, and a Random condition. Both blocks consisted of 225 trials, of which 25 were NoGo trials (i.e. digit ‘3’) and 200 were Go trials. Digits were presented for only 250ms, and participants had up to 900ms to make a response. The digits were presented in one of five randomly selected font sizes 48, 72, 95, 100, or 120 pixels). The measures of interest were the average RT for correct Go trials for the Ascending and Random conditions, the number of omission error (i.e., failure to respond on a Go trial), and the number of commission errors (i.e., incorrectly responding on a NoGo trial).

*Signal Detection task:* A Signal Detection task was used to assess perceptual decisions and probabilistic reward learning [18]. Due to a task coding error, the reward manipulation aspect of the task was not implemented as intended and therefore is not further described. We nonetheless chose to the analyze test-retest reliability of measures related to perceptual decision making because several elements of the task, notably very fast stimulus presentations, pressured responses, and long task duration, present unique challenges for remote and unsupervised cognitive testing in a patient population. In the task, participants viewed very brief presentations of cartoon smiley faces and were instructed to identify whether the length of the line for the smile was either “short” or “long”. On each trial, a mouth-less cartoon face was presented for 500ms following which either a short (11.5mm) or long mouth (13mm) appeared for 100ms. Participants used the ‘Z’ and ‘M’ keys to indicate if the smile was short or long, respectively. The task consisted of 3 blocks of 100 trials each, a fixation cross appeared for 500ms between trials. The order of long and short mouths was random, with each appearing 50 times within a block. Measures of interest included average RT for correct trials and one derived measure, discriminability, which is the ability for participants to discriminate between the long and short stimulus [18].

**Supplementary Results**

**Table S1**: Task performance with alternative measures of interest.

|  |  | Parkinson’s Patients | Older Adults |  |
| --- | --- | --- | --- | --- |
|  | Measure of interest | Mean performance at t1 | Mean performance at t1 | *p*-value |
| Trail Making | A – Number of errors | 0.30 (1.23) | 0.35 (0.88) | 0.82 |
|  | B – Number of errors | 2.04 (4.21) | 0.97 (2.0) | 0.11 |
| Stroop | Congruent – Accuracy (%) | 0.90 (0.20) | 0.96 (0.12) | 0.085 |
|  | Noncongruent – Accuracy (%) | 0.79 (0.22) | 0.88 (0.20) | 0.064 |
| SART | Ascending – Number of errors | 7.98 (9.16) | 9.31 (19.82) | 0.61 |
|  | Random – Number of errors | 9.55 (12.14) | 7.66 (11.76) | 0.31 |
| Signal Detection | Accuracy (%) | 0.81 (0.089) | 0.89 (0.085) | **<0.001** |

**Table S2**: Practice effects via paired t-tests

|  |  | Parkinson’s Patients | | | | Older Adults | |
| --- | --- | --- | --- | --- | --- | --- | --- |
|  | Measure of interest | Avg. change  (t2-t1) | t | *p-* value | Avg. change  (t2-t1) | t | *p*-value |
| Digit Span | Forward (# digits) | -0.52 | -2.57 | **0.012** | 0.054 | 0.20 | 0.85 |
|  | Reverse (# digits) | -0.41 | -2.37 | **0.021** | 1.00 | 3.61 | **<0.001** |
| Trail Making | A – Time to complete (sec) | 3.28 | 1.47 | 0.15 | 2.10 | 1.065 | 0.30 |
|  | B – Time to complete (sec) | -1.70 | -0.36 | 0.72 | -3.15 | -1.34 | 0.19 |
| Stroop | Congruent – RT (msec) | 39.50 | 2.30 | **0.025** | 20.75 | 1.08 | 0.29 |
|  | Noncongruent – RT (msec) | 56.72 | 1.61 | 0.11 | 23.96 | 0.83 | 0.41 |
|  | Stroop effect (Noncon-con) | 19.23 | 0.72 | 0.48 | 9.35 | 0.41 | 0.69 |
| SART Ascending | RT (msec) | -15.96 | -2.02 | **0.048** | -11.36 | -0.92 | 0.37 |
|  | Omission errors (# errors) | -0.63 | -0.33 | 0.75 | -5.72 | -1.52 | 0.14 |
|  | Commission errors (# errors) | -0.32 | -0.50 | 0.62 | -0.18 | -0.44 | 0.66 |
| SART Random | RT (msec) | -23.44 | -2.42 | **0.018** | -39.42 | -2.81 | **0.0087** |
|  | Omission errors (# errors) | -2.83 | -1.77 | 0.082 | -0.52 | -0.17 | 0.87 |
|  | Commission errors (# errors) | -0.61 | -1.03 | 0.31 | 0.97 | 1.49 | 0.15 |
| Signal Detection | RT (msec) | -10.13 | -1.44 | 0.16 | 3.58 | 0.67 | 0.51 |
|  | Discriminability | 0.28 | 4.30 | **<0.001** | 0.097 | 0.95 | 0.35 |

**Table S3:** Practice effects of alternative measures of interest via paired t tests

|  |  | Parkinson’s Patients | | | Older Adults | | |
| --- | --- | --- | --- | --- | --- | --- | --- |
|  | Measure of interest | Avg. change  (t2-t1) | t | *p*- value | Avg. change  (t2-t1) | t | *p*-value |
| Trail Making | A – Number of errors | 0.28 | 1.91 | 0.062 | -0.23 | -1.49 | 0.15 |
|  | B – Number of errors | 0.54 | 0.94 | 0.35 | -0.12 | -0.32 | 0.75 |
| Stroop | Congruent – Accuracy (%) | 0.027 | 0.89 | 0.38 | 0.010 | 0.48 | 0.63 |
|  | Noncongruent – Accuracy (%) | 0.0032 | 0.14 | 0.89 | 0.041 | 1.30 | 0.20 |
| SART | Ascending – Number of errors | -0.48 | -0.47 | 0.64 | -2.91 | -1.54 | 0.13 |
|  | Random – Number of errors | -1.73 | -2.01 | **0.047** | 0.25 | 0.17 | 0.87 |
| Signal Detection | Accuracy (%) | 0.034 | 3.81 | **<0.001** | 0.015 | 1.54 | 0.13 |

**Table S4**: Test-retest reliability of alternative measures of interest via ICCs

|  |  | Parkinson’s Patients | | | Older Adults | | |
| --- | --- | --- | --- | --- | --- | --- | --- |
|  | Measure of interest | ICC | *p*-value | 95% Confidence Interval | ICC | *p*-value | 95% Confidence Interval |
| Trail Making | A – Number of errors | 0.66 | **<0.001** | [0.47, 0.79] | 0.33 | **0.027** | [-0.003, 0.61] |
|  | B – Number of errors | 0.69 | **<0.001** | [0.53, 0.80] | 0.38 | **0.013** | [0.051, 0.64] |
| Stroop | Congruent – Accuracy (%) | 0.12 | 0.19 | [-0.14, 0.36] | 0.19 | 0.14 | [-0.15, 0.49] |
|  | Noncongruent – Accuracy (%) | 0.74 | **<0.001** | [0.59, 0.84] | 0.44 | **0.0045** | [0.12, 0.67] |
| SART | Ascending – Number of errors | 0.36 | **<0.001** | [0.20, 0.51] | 0.51 | **<0.001** | [0.31, 0.67] |
|  | Random – Number of errors | 0.60 | **<0.001** | [0.47, 0.70] | 0.48 | **<0.001** | [0.27, 0.65] |
| Signal Detection | Accuracy (%) | 0.75 | **<0.001** | [0.56, 0.86] | 0.60 | **<0.001** | [0.34, 0.77] |
